# Helmet Use Among E-Bike, Pedal Bike, and E-Scooter Riders in Canberra: A Cross-sectional Survey Study (Phase 4) [Protocol]

**DOI:** 10.64898/2026.03.04.26347651

**Authors:** Alan Silburn

## Abstract

**Background:** Helmet use is a proven safety measure that reduces the risk of head injury among cyclists and e-scooter riders. Despite legal requirements for pedal bikes and e-bikes in Australia, compliance varies, particularly among users of electric vehicles. The growing popularity of e-bikes and e-scooters in urban areas presents new public health challenges, yet observational data on helmet use, behavioural determinants, and the effectiveness of safety interventions remain limited.

**Aim:** Phase 4 of the Helmet Use in Canberra study aims to identify demographic and behavioural predictors of unsafe riding and to explore perceived barriers and facilitators to helmet use, including compliance with existing regulations.

**Methods:** A cross-sectional survey will be administered to Canberra residents aged 18 years or older, both online and in-person. The survey will assess attitudes toward helmet use, perceptions of head injury risk, and the deterrent effect of fines. Data will capture demographic characteristics, vehicle type, riding behaviours under varying conditions, and opinions regarding mandatory helmet laws and signage interventions. Survey responses will be de-identified, securely stored, and analysed using descriptive statistics and ordinal logistic regression to evaluate factors influencing compliance. Survey findings will be triangulated with observational and hospital data from earlier study phases.

**Expected Results:** The survey is anticipated to provide insights into public attitudes toward helmet use, the perceived effectiveness of fines as behavioural deterrents, and the acceptability of policy interventions. These findings will inform evidence-based strategies to improve helmet compliance and reduce head injuries among urban riders.

**Trial Registration:** Australian and New Zealand Clinical Trials Registry (ANZCTR) [ACTRN12626000245392].

## 1. SYNOPSIS

Helmet use is a well-established safety measure that significantly reduces the risk of head injury among cyclists and e-scooter riders. Despite Australian legislation mandating helmet use for pedal bikes and e-bikes, compliance remains variable, particularly among users of electric vehicles. The increasing popularity of e-bikes and e-scooters in urban environments presents new challenges for injury prevention. Understanding helmet-wearing behaviour, evaluating the impact of targeted interventions, and linking observational findings to hospital presentations are critical for informing evidence-based public safety policies.

This sub-study forms part of a larger study that employs a multi-method approach comprising three complementary components: a quasi-experimental observational study of helmet use in public settings, retrospective analysis of head injury presentations at The Canberra Hospital, and a cross-sectional survey assessing public attitudes toward helmet use and deterrent fines. Observational data will be collected across three high-traffic urban bike paths, with pre- and post-installation of signage emphasising either health benefits or legal penalties. Hospital data will provide context regarding injury outcomes associated with helmet use trends. Survey data will inform potential policy strategies to increase compliance.

The primary objective of the larger study is to assess helmet use rates among riders of e-bikes, pedal bikes, and e-scooters and to evaluate the effectiveness of signage interventions. Secondary objectives include examining demographic differences in helmet use, exploring associations with head injury presentations, and assessing public perceptions of deterrent fines. This protocol outlines study rationale, design, methodology, ethical considerations, and anticipated outcomes to ensure robust and meaningful findings for injury prevention in Canberra.

## 2. RATIONALE / BACKGROUND

Head injuries associated with cycling and micromobility vehicles represent a substantial public health concern, with consequences ranging from mild concussions to severe traumatic brain injury. Helmet use consistently mitigates the risk of head injury, with protective effects well documented across conventional bicycles, e-bikes, and e-scooters (1,2). For example, a meta-analysis of over 64,000 cyclist crash cases estimated that helmet use is associated with ∼51% lower risk of head injury and ∼69% lower risk of serious head injury compared with non-use, supporting widespread helmet use in cycling safety planning (1). Moreover, Høye’s review suggests that mandatory helmet legislation is associated with reductions of ∼20% in head injuries and ∼55% in serious head injuries (3). In the Australian context, the introduction of universal bicycle helmet laws in the early 1990s was followed by an immediate ∼46% decline in cycling fatality rates, though the effect on nonfatal head injuries is less consistently documented (4).

However, compliance remains inconsistent, particularly among users of electric mobility devices in urban areas. Observational studies in Australia show significant proportions of e-scooter riders not wearing helmets even where mandated (5). A study in Melbourne found that ∼50% of e-scooter injury presentations involved head injuries, while only one-third of riders reported wearing helmets (6). The rapid growth in e-scooter adoption has been mirrored by increasing hospital presentations for related injuries and calls for updated regulatory frameworks (7,8), highlighting the need to explore factors influencing helmet-wearing behaviour.

Previous observational and epidemiological studies suggest that behavioural, infrastructural, and policy factors influence helmet-wearing behaviour. Interventions such as signage emphasising health benefits or legal penalties may promote compliance, though evidence from Australian urban settings is still limited. Complementing observational data with hospital head injury records enables a triangulated view of how helmet behaviour maps to clinical outcomes, while survey data can shed light on the acceptability of enforcement strategies and what might drive behavioural change. In addition, legal context and enforcement levels may further shape compliance patterns.

Helmet laws and fines vary widely across Australian jurisdictions, which may influence helmet-wearing behaviour. In the ACT, riders face a fine of AUD 121 for not wearing a helmet, comparatively modest next to AUD 344 in New South Wales and Tasmania, AUD 227 in Victoria, and AUD 205 in South Australia. Fines are lower in Queensland (AUD 137), Western Australia (AUD 50), and the Northern Territory (AUD 25) (9). Enforcement intensity also differs, with NSW police reportedly issuing ∼500 fines per month (9). These variations suggest that both fine magnitude and policing practices may shape helmet-wearing behaviour, providing critical context for interpreting compliance trends in Canberra and for designing targeted interventions such as signage emphasising health benefits or legal penalties.

Thus, linking helmet use behaviour, signage-based interventions, and hospital head injury trends offers a promising approach to inform effective local injury prevention policy in Canberra.

## 3. AIMS / OBJECTIVES / HYPOTHESES

### 3.1. Aim

Phase 3 aims to identify demographic and behavioural predictors of unsafe riding and explore perceived barriers and facilitators to helmet use and compliance with regulations.

### 3.2. Primary Objectives

- To examine attitudes to helmet use by age group, gender, vehicle type, and environmental conditions.

### 3.3. Secondary Objectives

- To identify demographic and contextual predictors of helmet compliance.

### 3.4. Hypotheses

- Perceptions of helmet utility will vary by age, gender, and vehicle type.

## 4. STUDY PHASE

- Phase 4: Cross-Sectional Survey

A voluntary survey of Canberra residents aged 18 years or older will capture attitudes toward helmet use and perceptions of deterrent fines. Survey data will complement observational and hospital findings by providing insight into behavioural drivers, policy acceptability, and potential intervention strategies.

The survey details and variables examined are further detailed in Appendix 1: Survey Outline.

## 5. RESEARCH PLAN / STUDY DESIGN

### 5.1. Data Sources / Collection

#### 5.1.1. Cross-Sectional Survey

A voluntary survey will be administered online and in-person to Canberra residents aged 18 years or older. The survey will assess attitudes toward helmet use, perceived risks of non-compliance, and the potential deterrent effect of fines. Participation will be voluntary, with informed consent obtained prior to completion. Responses will be collected anonymously and stored securely.

#### 5.1.2. Data Management and Security

Survey responses will be stored on secure, password-protected institutional servers accessible only to authorised personnel. Following UTAS IT Services’ recommendation, a UTAS Dropbox will be utilised. Raw data will not be shared externally. Data management procedures will comply with institutional, NHMRC, and local privacy guidelines, with routine backups and secure retention for a minimum of five years.

#### 5.1.3. Quality Control and Fidelity Monitoring

Coding will be conducted by trained researchers to ensure reliability. Survey instruments and data extraction protocols will undergo pilot testing to confirm clarity, completeness, and accuracy. Regular review meetings will monitor data integrity, adherence to protocol, and timely resolution of any discrepancies.

### 5.2. Population / Sample Size

#### 5.2.1. Population

Participants will be adults aged 18 years and older who have used a bike or scooter in Canberra in the past 12 months.

#### 5.2.2. Sample Size

For the survey component, a minimum of 300 respondents is targeted to provide sufficient power for ordinal logistic regression analyses and to ensure demographic representativeness.

### 5.3. Statistical Analyses

All statistical analyses will be conducted in R®. Multiple imputation will address missing data where appropriate. Sensitivity analyses will include alternative specifications of period effects, adjustment for baseline covariates, per-protocol analyses limited to high-fidelity clusters, and subgroup analyses by age and baseline compliance.

#### 5.3.1. Survey Data

Descriptive statistics will summarise participant attitudes toward helmet use and perceptions of fines. Ordinal logistic regression models will evaluate which fine amounts are most likely to influence compliance, controlling for demographic factors and prior helmet-wearing behaviour. Integration of survey findings with observational and hospital data will allow triangulation of behavioural, attitudinal, and injury outcome evidence.

## 6. ETHICAL CONSIDERATIONS

### 6.1. Recruitment and Selection of Participants

Recruitment will be obtained using voluntary participation. The survey will be linked via QR code to informative posters displayed at various locations throughout Canberra, including hospitals, universities, community centres, bike/scooter retailers, and known bike/scooter hire locations.

Participants will be adults aged 18 years and older who have used a bike or scooter in Canberra in the past 12 months. There will be no exclusion based on gender, ethnicity, or socioeconomic status. Participants must be able to read English and provide informed consent.

### 6.2. Informed Consent

Informed consent will be obtained electronically before survey commencement. A link to the study protocol and ANZCTR site will be provided for transparency regarding the study purpose, procedures, risks, benefits, confidentiality measures, and the voluntary nature of participation. Participants will be required to confirm consent before accessing survey questions. Participants may withdraw at any time before submission; once submitted, responses cannot be withdrawn due to anonymity.

### 6.3. Confidentiality and Privacy

All data will be de-identified. All data will be stored securely on encrypted servers accessible only to authorised personnel. Any external sharing for publication or collaboration will be fully anonymised and aggregated.

### 6.4. Data Access and Dissemination

Only study investigators and authorised data managers will access identifiable data (e.g., electronic medical records). Results will be disseminated at the aggregate level to policymakers, urban planners, cycling advocacy groups, and academic audiences.

### 6.5. Support for Staff

All staff and participants will receive contact details for the research team and information on support services (e.g., Lifeline, Beyond Blue) if discussions of cycling injuries cause distress.

### 6.6. Aboriginal and Torres Strait Islander Data

The study does not seek to specifically identify Aboriginal or Torres Strait Islander children. Should the research team wish to analyse outcomes by Indigenous status, this will only occur with explicit consent and following approval from an Aboriginal Human Research Ethics Committee (HREC), in line with the NHMRC Ethical Conduct in Research with Aboriginal and Torres Strait Islander Peoples and communities (2018). The protocol will be amended and resubmitted for additional review if such analyses are proposed.

### 6.7. Data Storage and Record Retention

Electronic data will be stored on encrypted institutional servers, with routine backups maintained according to institutional and NHMRC guidelines. All data will be retained for a minimum of five years following study completion, after which secure deletion procedures will be implemented. Hard copies, if generated, will be securely shredded after the retention period. Data management plans will ensure compliance with applicable regulations, including the NSW Health Privacy Manual and NHMRC National Statement on Ethical Conduct in Human Research.

### 6.11. Ethics Approval and Oversight

Ethics approval will be sought from the University of Tasmania HREC before commencement. If Indigenous data collection is proposed, additional review will be sought from an Aboriginal HREC. All applications will be submitted and approved before any participant recruitment or data collection begins. The study protocol will be registered in the Australian and New Zealand Clinical Trials Registry (ANZCTR), and the registration details will be publicly available prior to study commencement. Publication of the protocol will also occur before the start of the study to ensure transparency and facilitate reproducibility. Continuous oversight of the study will be maintained throughout its duration, including monitoring participant safety, data integrity, and adherence to the approved protocol. Any adverse events will be promptly documented and reported to the HREC and relevant authorities, and participants will be informed of any developments that may impact their continued participation. Amendments to study procedures will be submitted for ethics review and approval before implementation.

## Supporting information

Phase 4_Participant Information Sheet

Phase 4_Appendix 1. Survey Outline

Phase 4_Poster

**LIST OF INVESTIGATORS AND PARTICIPATING INSTITUTIONS**

**Chief Investigator:**

Dr Brenton Systermans

Course Coordinator & Senior Lecturer, Healthcare in Remote and Extreme Environments,Tasmanian School of Medicine, University of Tasmania.

**Principal Investigator:**

Mr Alan Silburn

Paramedic & Registered Nurse (Division 2), NSW Ambulance / NSW Health

Academic, Western Sydney University

Fellow of the Royal Society for Public Health (FRSPH)

**Associate Investigator:**

Dr Sean Chan

Intensive Care Staff Specialist, The Canberra Hospital

State Medical Director, Donate Life ACT

Deputy Director, ACT Trauma Service

**Associate Investigator:**

Dr Thomas Georgeson

Emergency Staff Specialist, The Canberra Hospital

**Participating Institutions:**

University of Tasmania

Churchill Ave, Dynnyrne TAS 7005, Australia

The Canberra Hospital

Yamba Dr, Garran ACT 2605

## 7. OUTCOMES AND SIGNIFICANCE

The primary outcome of this study will define public attitudes toward helmet use and the perceived effectiveness of fines as behavioural deterrents, thereby informing feasible and acceptable policy interventions.

The significance of this study is reinforced when considered as part of the larger study. By integrating observational, clinical, and attitudinal data, it will generate a robust, context-specific evidence base for designing interventions aimed at improving helmet compliance among urban riders. Insights from this study may inform policy decisions, including the optimal deployment of educational signage, the calibration of deterrent fines, and broader urban planning strategies to enhance rider safety.

At a population level, the findings have the potential to reduce the incidence and severity of cycling and e-scooter-related head injuries, decrease healthcare utilisation, and promote a culture of safety in urban mobility. The evidence generated will be directly relevant to local policymakers, health authorities, transport planners, and advocacy organisations, supporting the development of scalable, cost-effective, and socially acceptable strategies to enhance helmet use. Ultimately, the study contributes to a broader understanding of behavioural determinants of safety compliance in rapidly evolving urban transport contexts and may serve as a model for similar initiatives in other Australian cities.

## 8. TIMELINES / MILESTONES

Months 0–2: Ethics approval, survey design and pilot testing.

Months 2–4: Survey distribution.

Months 4–6: Data cleaning, coding, and analysis.

Months 6–12: Dissemination of results to stakeholders, manuscript preparation, and policy recommendations.

## 9. PUBLICATION POLICY

Findings will be disseminated via peer-reviewed publications, conference presentations, and reports to policymakers, urban planners, and advocacy organisations. Authorship will adhere to ICMJE guidelines.

## Declarations

### Abbreviations

Not applicable

### Human Ethics

Ethics approval has been obtained from the University of Tasmania Human Research Ethics Committee (HREC) [H40545].

### Registration

The Helmet Use in Canberra study has been registered in the Australian and New Zealand Clinical Trials Registry (ANZCTR) [ACTRN12626000245392].

### Consent for publication

The author consents to the publication of this protocol.

### Availability of data and materials

Available at request from the corresponding author.

### Competing Interests

The author declares that they have no known competing interests or personal relationships that could have appeared to influence the work reported in this paper.

### Funding

The author declares that they did not receive funding for this article.

### Authors’ contributions

The author solely contributed to the conception, design, analysis, and drafting of the manuscript.

## Acknowledgements

Not applicable

## Notes

### Competing Interest Statement

The authors have declared no competing interest.

### Clinical Trial

ACTRN12626000245392

### Funding Statement

This study did not receive any funding.

### Author Declarations

This study has been approved by the University of Tasmania Human Research Ethics Committee [H40545].

