## Supplementary material for "Helmet Use Among E-Bike, Pedal Bike, and E-Scooter Riders in Canberra: A Cross-sectional Survey Study (Phase 4) [Protocol]": Phase 4_Participant Information Sheet

### ANZCTR Registration: ACTRN12626000245392

**Alan Silburn GradDipCH GradDipHREE MPH FAWM FAEEM FRSPH[1]***

1 University of Tasmania, Dynnyrne TAS 7005, Australia
Tele: 0449 107 944

#### STUDY ABSTRACT

**Background:** Helmet use is a proven safety measure that reduces the risk of head injury among cyclists and e-scooter riders. Despite legal requirements for pedal bikes and e-bikes in Australia, compliance varies, particularly among users of electric vehicles. The growing popularity of e-bikes and e-scooters in urban areas presents new public health challenges, yet observational data on helmet use, behavioural determinants, and the effectiveness of safety interventions remain limited.

**Aim:** Phase 4 of the Helmet Use in Canberra study aims to identify demographic and behavioural predictors of unsafe riding and to explore perceived barriers and facilitators to helmet use, including compliance with existing regulations.

**Methods:** A cross-sectional survey will be administered to Canberra residents aged 18 years or older, both online and in-person. The survey will assess attitudes toward helmet use, perceptions of head injury risk, and the deterrent effect of fines. Data will capture demographic characteristics, vehicle type, riding behaviours under varying conditions, and opinions regarding mandatory helmet laws and signage interventions. Survey responses will be de-identified, securely stored, and analysed using descriptive statistics and ordinal logistic regression to evaluate factors influencing compliance. Survey findings will be triangulated with observational and hospital data from earlier study phases.

**Expected Results**: The survey is anticipated to provide insights into public attitudes toward helmet use, the perceived effectiveness of fines as behavioural deterrents, and the acceptability of policy interventions. These findings will inform evidence-based strategies to improve helmet compliance and reduce head injuries among urban riders.

---

**1. Who is conducting this research?**

This study is being conducted by researchers from the University of Tasmania.

**Chief Investigator:**

Dr Brenton Systermans

Course Coordinator & Senior Lecturer, Healthcare in Remote and Extreme Environments,

Tasmanian School of Medicine, University of Tasmania.

**Principal Investigator:**

Mr Alan Silburn

Paramedic & Registered Nurse (Division 2), NSW Ambulance / NSW Health

Academic, Western Sydney University

Fellow of the Royal Society for Public Health (FRSPH).

This study has been registered with the Australian and New Zealand Clinical Trials Registry [ACTRN12626000245392], and ethics approval was sought from the University of Tasmania Human Research Ethics Committee (HREC) [H40545] before recruitment began.

**2. What is the purpose of the study?**

It is argued that helmet use significantly reduces the risk of head injury for riders of bicycles, e-bikes, and e-scooters. Although helmet use is legally required in the ACT, not all riders comply.

This survey aims to better understand:

- Attitudes toward helmet use
- Perceived risks of riding without a helmet
- Whether fines act as a deterrent
- Barriers and facilitators to helmet compliance

Your responses will help inform evidence-based strategies to improve rider safety in Canberra.

**3. Who can participate?**

You may participate if you:

- Are aged 18 years or older; and
- Have ridden a bicycle, e-bike, or e-scooter in Canberra in the past 12 months; and
- Can read and understand English.

Participation is entirely voluntary.

**4. What does participation involve?**

If you agree to participate, you will complete an anonymous survey online (via QR code).

The survey will take approximately 5-10 minutes to complete and includes questions about:

- Your age group and general demographics
- The type of vehicle you ride
- Your helmet-wearing habits
- Your views on helmet laws and fines
- Situations where you may or may not wear a helmet

You may skip any question you do not wish to answer. You may withdraw at any time before submitting the survey. Once submitted, responses cannot be withdrawn because they are anonymous and cannot be linked back to you.

**5. Are there any risks?**

This study is low risk. Some participants may feel mild discomfort when thinking about cycling injuries. If participation raises any concerns for you, you may contact support services such as Lifeline (13 11 14) or Beyond Blue (1300 22 4636).

**6. Are there any benefits?**

There are no direct personal benefits. However, your participation will contribute to research that may improve urban transport safety, inform public policy, and reduce head injuries in Canberra.

**7. Privacy and confidentiality**

Your survey responses are completely anonymous. No identifying information will be collected.

All data will be stored securely on password-protected institutional servers and accessible only to authorised research personnel. Data will be retained for a minimum of five years in accordance with national research guidelines and then securely deleted.

Results will be reported in aggregate form (for example, as grouped statistics) in academic publications and reports. Individual participants will not be identifiable.

**8. Do I have to take part?**

No. Participation is entirely voluntary. Choosing not to participate will have no consequences for you. Completing and submitting the survey indicates your consent to participate.

**9. Further information**

If you have questions about the study, please contact:

Mr Alan Silburn


This study has been approved by the University of Tasmania Human Research Ethics Committee. If you have concerns or complaints about the conduct of this study, you can contact the Executive Officer of the HREC on (03) 6226 6254 or. The Executive Officer is the person nominated to receive complaints from research participants. You will need to quote H40545.
