## Supplementary material for "Helmet Use Among E-Bike, Pedal Bike, and E-Scooter Riders in Canberra: A Cross-sectional Survey Study (Phase 4) [Protocol]": Phase 4_Appendix 1. Survey Outline

**Alan Silburn GradDipCH GradDipHREE MPH FAWM FAEEM FRSPH[1*]**

1 University of Tasmania, Dynnyrne TAS 7005, Australia
Tele: 0449 107 944

The survey will be conducted through Qualtrics and is available from the following link:

https://qualtricsxm72ynsrj3c.qualtrics.com/jfe/form/SV_4ZymHgdbjWRGCZU

**Survey Outline**

Following QR code scanning -

The landing page will include the UTAS logo and the following statement.

"This survey is targeted towards Canberra residents, aged 18 years or older, and aims to capture attitudes toward helmet use, health benefits, and perceptions of deterrent fines."

Further information is available from Alan Silburn

Trial Registration: (https://anzctr.org.au/ACTRN12626000245392.aspx.)

Study Protocol (doi link)

Participant Information Sheet (doi link)

"Consent will be implied through your submission of the survey, indicating that you have read the information provided and agree to participate."

START SURVEY button -

The subsequent survey questions will include variables such as:

- Age Group (18-24, 25-29, 30-34, 35-39, 40-44, 45-50, 50+)
- Gender (Male, Female, Transperson, Prefer not to say)
- Vehicle type (E-bike, Pedal bike, E-scooter, Pedal scooter)
- Reason for travel (Work, Recreation, Primary transport)
- Do you wear a helmet when riding your personal vehicle (Never, Sometimes, Often, Always)
- Do you wear a helmet when riding a hire vehicle (Never, Sometimes, Often, Always)

- At night, do you wear a helmet (Never, Less often compared to day, More often compared to day, Always)
- If the weather is poor (such as raining), do you wear a helmet (Never, Less often compared to fine weather, More often compared to fine weather, Always)
- After consuming alcohol, do you wear a helmet (Never, Less often compared to no alcohol, More often compared to no alcohol, Always)
- Should helmets be mandatory for adults (Yes/No)
- Should helmets be mandatory for children (<18yr) (Yes/No)
- Do you think helmets reduce head injuries during accidents (Yes/No)
- Do you think public signs about the health benefits of wearing helmets are/would be beneficial (Yes/No)
- Do you think public signs about the penalties for not wearing helmets are/would be beneficial (Yes/No)
- What amount of fine would be a deterrent for you ($100, $200, $300, $400, $500+)

SUBMIT SURVEY button –

UTAS logo and “Thankyou for your contribution” message.
