## Supplementary material for "Helmet Use Among E-Bike, Pedal Bike, and E-Scooter Riders in Canberra: A Cross-sectional Survey Study (Phase 4) [Protocol]": Phase 4_Poster

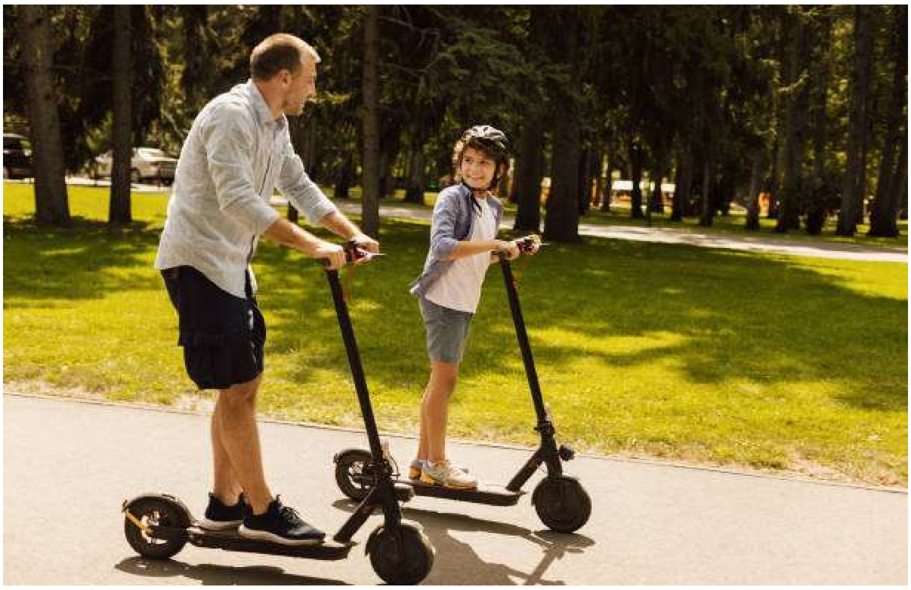

### SHOULD HELMETS BE MANDATORY?
### DO YOU THINK THEY HELP?

We are inviting Canberra residents aged 18 years and over to take part in a short **multiple-choice** survey about helmet use when riding bikes and e-scooters.
The survey can be completed using the QR code and asks about when you wear a helmet, your views on helmet safety, and what encourages helmet wearing. We will also ask general questions about your age group, the type of bike or scooter you use, your riding habits in different situations, and your opinions on helmet laws and safety signage.

All responses are anonymous and confidential. Your answers will be securely stored and analysed to better understand what influences helmet use in Canberra and how rider safety can be improved.

This study is registered with the Australian and New Zealand Clinical Trials Registry [ACTRN12626000245392] and the University of Tasmania Ethics Committee [H40545].

#### Survey Responses Needed

**We want to know why riders DO and DON'T wear helmets in the ACT.**

**Should children have to wear helmets?**

**Please scan the QR code and fill out the 15-question survey.**

Further information is available from Alan Silburn

UNIVERSITY of TASMANIA

QR code
